# Rapid realist review: Anxiolytic effects of music therapy on mechanically ventilated patients

**DOI:** 10.1101/2021.09.11.21263390

**Authors:** Hannah Watson, Patrick Marshall

## Abstract

**Aims:** To review the literature focusing on anxiety and music therapy as a non-pharmacological anxiolytic for patients receiving mechanical ventilation and to determine the contexts, mechanisms and outcomes of what works for whom and under what circumstances.

**Background:** Mechanical ventilation is one of the numerous processes that a critically ill patient requiring single or multi organ support may experience. This is often frightening and perplexing for the patient, particularly when mechanically ventilated. The abundance of interventions, monitoring and unfamiliar noises can precipitate feelings of stress and anxiety which is common within this population of patients leading to prolonged hospital stay and increases in mortality and morbidity.

**Methods:** A rapid realist review (Abrahamson *et al*. 2020) was undertaken applying the realist methodology to a search of the literature using CINHAL, MEDLINE and Cochrane library, PsycIFNO, PubMed and EMBASE along with searching for the grey literature using an experience library technician, Google Scholar, Google, OpenGrey and the British Library ETHOS.

**Results:** Twenty-one studies were included in the review. From the heterogeneity amongst the literature and the poor quality of evidence it was ascertained what worked for whom and under what circumstances. No negative outcomes for patients were noted in the review thus suggesting that music may have a place within critical care to help reduce anxiety. Given the heterogeneity of the evidence there is scope to review this topic further.

**Conclusion:** The overarching conclusion was that music could help reduce anxiety in the critically ill mechanically ventilated patient. Thematic analysis helped identify what quantifies the markers of anxiety; furthermore, it noted alternative themes that could be explored through more research. Theories could be developed and implemented into a protocol for practice; however, it would be based on the researchers’ own experience due to the poor-quality heterogeneous evidence.

## BACKGROUND

The abundance of noise, frequent interventions, machinery and monitoring precipitates a frightening and perplexing environment for critically ill patients (Mpouzika *et al*. 2017). Feelings of psychoemotional distress and anxiety are common amongst this population of patients and this can lead to prolonged hospital stay, mortality and morbidity (Chlan *et al*. 2013a; Khalaiha *et al*. 2011; *Bradt et al*. 2010; Chan *et al*. 2008; Almured & Petersson, 2003). Alasad *et al*. (2015) note that patients in ICU can recall the positives and negative aspects of their stay; however, most recall the most unpleasant aspects, which can lead to posttraumatic stress disorder (PTSD) as noted by Chahraoui *et al*. (2015) and Wu *et al*. (2018) noting that a lack of emotional support and the perceived worsening of life and future uncertainty during a patients stay in the Intensive Care Unit (ICU) are a risk factor for posttraumatic stress, anxiety and depression (Barr *et al*. 2013). There is scope to assess patients’ memories, experiences and coping strategies post their stay in ICU; however, it is beyond the scope of this review to evaluate this.

Caine (2003) notes there are links between psychological stress and hypertension, hypotension, pain, cardiac disorders, respiratory pathologies, infection and tachycardia. Papathanassoglou *et al*. (2010) recognises that psychological stress heightens the pathophysiological sequlae through the release of neuropeptides, which can be measured to assess the stress and effectiveness of psychological support interventions particularly in non-communicative critically ill patients.

Bradt and Dileo (2014) suggested that music therapy may be beneficial for mechanically ventilated patients and noted a reduction in physiological measurements and anxiety in their Cochrane review, which was also noted in other systematic reviews by Umbrello, *et al* (2019), Davis and Jones (2012), Nilsson, (2008) and Biley (2000).

Sepsis, acute respiratory failure, cardiac failure, pneumonia, trauma and neuromuscular disease are amongst the many pathologies that the critically ill patient often endures requiring single or multiple organ support in particular; mechanical ventilation (Longmore *et al*. 2014; Loftus, 2010; Waldmann *et al*. 2008; Cooper *et al*. 2006). Patients report that the most frustrating part of being mechanically ventilated is the inability to communicate; followed by thirst, difficulty swallowing and pain (Flinterud & Anderson, 2015; Karlsson *et al*. 2012; Khalaia *et al*. 2011). These frustrations can lead to a range of emotions as reported by patients including: panic; fear; anger; helplessness; hopelessness. They report that they feel a loss of control and being overwhelmed in addition to being critically ill (Flinterud & Anderson, 2015; Karlsson *et al*. 2012; Khalaia *et al*. 2011).

This causes pain, anxiety and delirium, particularly in mechanically ventilated patients as they often require pharmacological management to aid with ventilator synchronisation and manage pain, which can be associated with adverse outcomes including prolonged hospital stay, ventilation and further complications increasing morbidity and mortality (Reade & Finfer, 2014; Barr *et al*. 2013 Khalaia *et al*. 2011; Thomas, 2003).

It is common for patients to undergo biological stress, physiological stress and psychological stress; particularly, those undergoing medical interventions (Mpouziki *et al*. 2017; Salamon *et al*. 2003). Salamon *et al*. (2003) notes that when haemostasis within a given organism is adjusted secondary to stimuli, this can be classified as biological stress. Allostatic loading (multiple causes of stress) can be pathological if not relieved (Clark *et al*. 2007; Salamon *et al*. 2003). Psychological phenomenon of stress is identified through feelings of apprehension, nervousness, anxiety, helplessness (Mpouziki *et al*. 2017; Salamon *et al*. 2003). Physiological stress is marked by hypertension, tachycardia, hyperventilation which are linked with ischaemia, fluctuations of body temperature, decreased appetite, enlarged pupils, pain and increased cortisol levels (Mpouzika *et al*. 2017; Clark *et al*. 2007; Salamon *et al*. 2003).

### Music and healing

Music has been recognised for its healing properties for centuries, resulting in positive impacts on human health and well-being throughout history from, ancient Greece through the Christian era, the renaissance, Romanticism and into the 20^th^ century where music therapy was recognised as a professional health care discipline (Davis & Jones, 2012; Carroll, 2011; Kemper & Danhauer, 2005, Cardozo, 2004; Darnley-Smith & Patey, 2003; Biley, 1999). Music has been linked with reducing anxiety, alleviating pain amongst dementia patients, psychiatric patients, patients suffering with anxiety and depression, burns patients, anaesthesiology, patients undergoing biopsy and end of life care (Li *et al*. 2017; Song *et al*. 2017; Atiwannapat *et al*. 2016; Shuman *et al*. 2016; Gutierrez & Camarena, 2015; Tanka & Nogawa, 2015; Brown *et al*. 2015; Matsota *et al*. 2013; Crawford *et al*. 2013; Jiang *et al*. 2013; To *et al*. 2013; Wakim *et al*. 2010; Kemper & Danhauer, 2005). Davis and Jones (2012) note that in 1978 music therapy was classified as the controlled use of music to influence the psychological, physiological and emotional experiences of patients during therapy and rehabilitation.

Music is complex as the mechanisms to how it works is unclear, the thalamus receives musical stimuli of which the effects are transmitted via the reticular activating system (RAS) towards the cerebrum stimulating imagination, intellect, memory and the automatic nervous system (O’Sullivan, 1991). Relaxation, meditation or hypnosis generates alpha waves in the brain indicating the brain is also affected my musical stimulus (O’Sullivan, 1991). Nitric oxide (NO) as a neurotransmitter has a role within the mechanisms of relaxation as it mediates the neuronal pathway from the auditory nerve to the cerebral cortex (Hall, 2016; Lindman & Chakinala, 2010; Salamon *et al*. 2003; Galley, 2000). NO as a lipophilic gas is released from the endothelial cells in response to chemical and physical stimuli by relaxing the blood vessels causing vasodilation thus facilitating in lowering blood pressure (Hall, 2016; Lindman & Chakinala, 2010; Salamon *et al*. 2003; Galley, 2000).

The complexity of human auditory and visual perception, and its link to intelligence are utilised to organise sound in the language self-expression and communication (Carroll, 2011). There are variations of genres of music and its benefits towards the listener (O’Sullivan, 1991). Cultural differences, associations and age offer a wide array of music to suit various people that can trigger positive or negative memories (Kemper & Danhauer, 2005; O’Sullivan, 1991). O’Sullivan (1991) notes that the tempo of music has a significant effect on one’s psychological state. Steady rhythms with a tempo of 60-70 beats per minute, low frequency tones and a relaxing melody are found to be soothing and linked with a lower heart rate and respiratory rate (Kemper & Danhauer, 2005; O’Sullivan, 1991). Loud music with a faster rhythm is associated with increased tensions, pain and anxiety; however, it is pertinent to note that music choice should be adapted to the listener’s needs and desires (Kemper & Danhauer, 2005; O’Sullivan, 1991).

### Aims and objectives

The aim of this review was:

- To review the available literature and search for the context, mechanisms and outcomes
- To ascertain the working mechanisms of the intervention
- To examine the theories surrounding the intervention
- To understand how the intervention can be translated into practice and understand the impact for patients

The population, intervention, comparison and outcome (PICO) guide was used to clarify the research questions (Aveyard, 2014):

#### Population

The review will focus on mechanically ventilated patients within an intensive care setting; however, for the purpose of theory development studies outside of a critical care setting may be included

#### Intervention

Music therapy as an adjunct to relieving stress

#### Comparison

Noise reducing headphones and usual intensive care therapy

#### Outcome

The psychological effects, behavioural effects, neurological effects and physiological effects of the patient will be the focus.

<u>Research questions:</u>

1. What are the key characteristics of music therapy that help to reduce anxiety? And
2. Does music therapy reduce anxiety in mechanically ventilated patients? And
3. How can this be implemented into practice?
4. What does this mean for patients?

## PRISMA

Meta-analysis and systematic reviews use the Preferred Reporting Items for Systematic Review and Meta-analysis (PRISMA) flow diagram as an evidence-based tool to demonstrate the inclusion and exclusion of articles used. Specifically systematic reviews use the PRISMA for the critical evaluation of a variety of published research (Moher *et al*, 2009; Liberati *et al*, 2009). (Refer to diagram 1). Initially this review was undertaken in 2019 therefore an updated search of the literature was carried out on 8^th^ July 2021 to ensure no articles were excluded from this review. Four additional articles were sourced one of which was included for analysis.

## METHODOLOGY

The clinical experience of working in intensive care settings offer a useful insight for investigating music therapy as an intervention as the overall effect of music is complex. Measuring the physiological effects alone is quantifiable; however, the patient’s experiences, feelings and thoughts towards the intervention offer valuable insight into the overall data extraction.

Humans are complex and unpredictable and to shed some light on music therapy as an intervention; mixed data and narrative data has offered multiple viewpoints on music as a phenomenon (Polit & Beck, 2006; Parahoo, 1997). A realist perspective was considered appropriate due to the complexity of the intervention, as the realist seeks to unpick the mechanisms of how the complex programmes/interventions succeed or fail, taking into account the contexts and settings. This review seeks to explore the available evidence rather than to generalise from the research; however, for the purpose of this review the literature has been critiqued to provide examples of the range of evidence available (Wong *et al*. 2013; Pawson *et al*. 2006; Pawson *et al*. 2004). Furthermore it was considered feasible to follow the rapid realist review (RRR) approach due to time constraints and limited resources (Saul *et al*. 2013).

### Rapid realist review (RRR)

Due to the substantial amount of investment over time a realist review requires which do not always meet the demands of the usual time restricted policy; the RRR is a tool that has been developed to apply a realist approach to a time-sensitive knowledge synthesis process to provide policy makers with a policy that identifies the knowledge and gaps within the literature identifying the CMO configurations in keeping with the core elements of realist philosophy (Wong *et al*. 2013; Saul *et al*. 2013; Pawson *et al*. 2004). This is a streamlined approach that avoids scope creep, which can be an identifiable risk factor for a realist synthesis (Saul *et al*. 2013). Although the process is like the Cochrane or systematic reviews, RRR is iterative in nature and the initial developments, search strategy, question development and quality approval may be revisited iteratively throughout the RRR process (Saul *et al*. 2013). RRR is limited as the rapid approach process may miss certain resources and references, which may introduce bias; however, this can be mitigated by using an expert panel or a local reference group ensuring critical elements are not missed (Saul *et al*. 2013).

### Ethics

Secondary data from established resources is used for this review, therefore did not require ethical approval as it did not involve people taking part in research indirectly and directly.

### Data gathering

This study used a comprehensive literature review; a preliminary search was conducted via search engines using key words to assess the scope and availability of the literature. Further iterative searching assessed the quality and quantity of the literature and identified further key words to be used in the more comprehensive search strategy.

### Search strategy

CINHAL, MEDLINE, Cochrane library, PsycINFO, PubMed and EMBASE were used to identify published literature along with searching for the grey literature using an experienced library technician, Google Scholar, Google, OpenGrey and the British Library ETHOS.

Boolean logic was applied when identifying key words during the search strategy (Aveyard, 2014): (music therapy* OR music intervention*) AND (critical care OR intensive care OR icu OR ITU) AND (anxiety OR worry OR stress OR fear* OR apprehension OR agitation OR tension)

### Inclusion/exclusion

It is not absolutely necessary in realist synthesis to exclude articles based on their study design, therefore; methodological filters have not been used in this review. Strict inclusion/exclusion criteria are also not a feature of a realist review as searching is iterative because new theories develop in light of emerging data, therefore; inclusion criteria are refined throughout the process of the review (Wong *et al*, 2013; Pawson *et al*, 2004). However, an inclusion/exclusion was applied to this review as follows:

> Adults (>18yrs), critically ill patients, ventilated patients, intensive care, ITU, ICU, music therapy, English articles, randomised controlled trials (RCT’s), cohort studies, Cochrane reviews, pilot studies, interviews and a mixed methods approach were all included in this review (n=27).
>
> Neonates and children (<18yrs), non-English articles, most articles focusing on non-critically ill patients (if the article focused on anxiety and music therapy in a non-critical ill setting then it was included) and music therapy that focused on relieving pain only were excluded.

Limited time and resources did not enable non-English articles to be included. The focus of this review is adults in critical care and the anxiolytic effect of music therefore children; neonates and non-critically ill patients were excluded to keep this review focused. There is scope to review pain and music therapy as it is linked with anxiety, however the focal point of this review is anxiety due to its subjective nature

Following the search strategy, the research questions were identified using the PICO analysis. Following the scope of the literature common themes were identified which led to further literature searching and further reading which narrowed the number of articles to 21 to be included in this review.

### Data extraction

The realist approach is theory driven to answer multiple questions and through identifying CMO configurations common themes were identified in the literature (Wong *et al*. 2013; Saul *et al*. 2013 Pawson *et al*. 2006; Pawson *et al*. 2004; Pawson & Tilley, 1997).

## RESULTS

The realist approach identifies theories within the mechanisms to delineate the interactions within the context. Refer to Table 1 for a detailed version of CMO configurations and Table 2 for a quick extraction of the data.

**Table 1:**
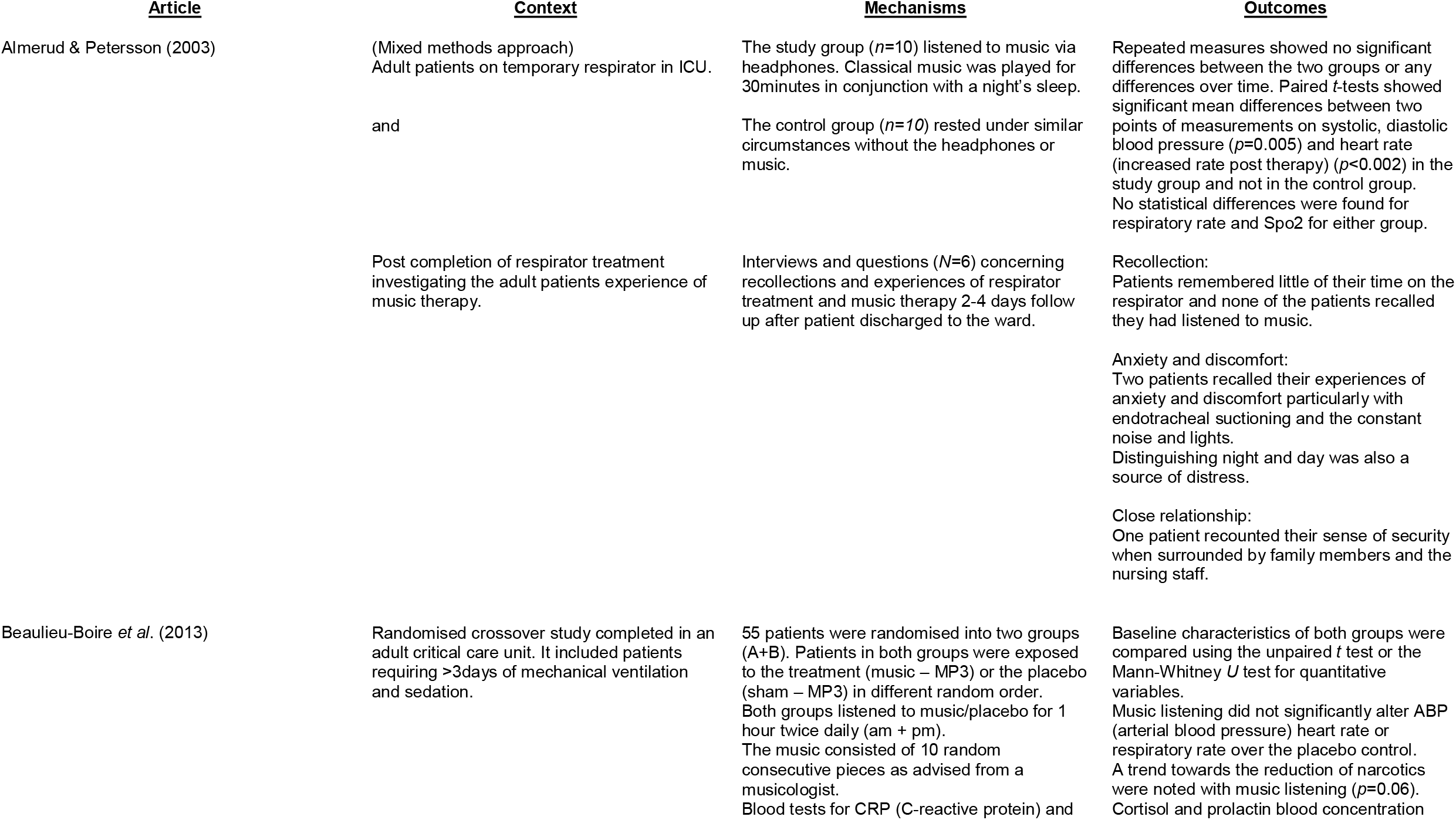
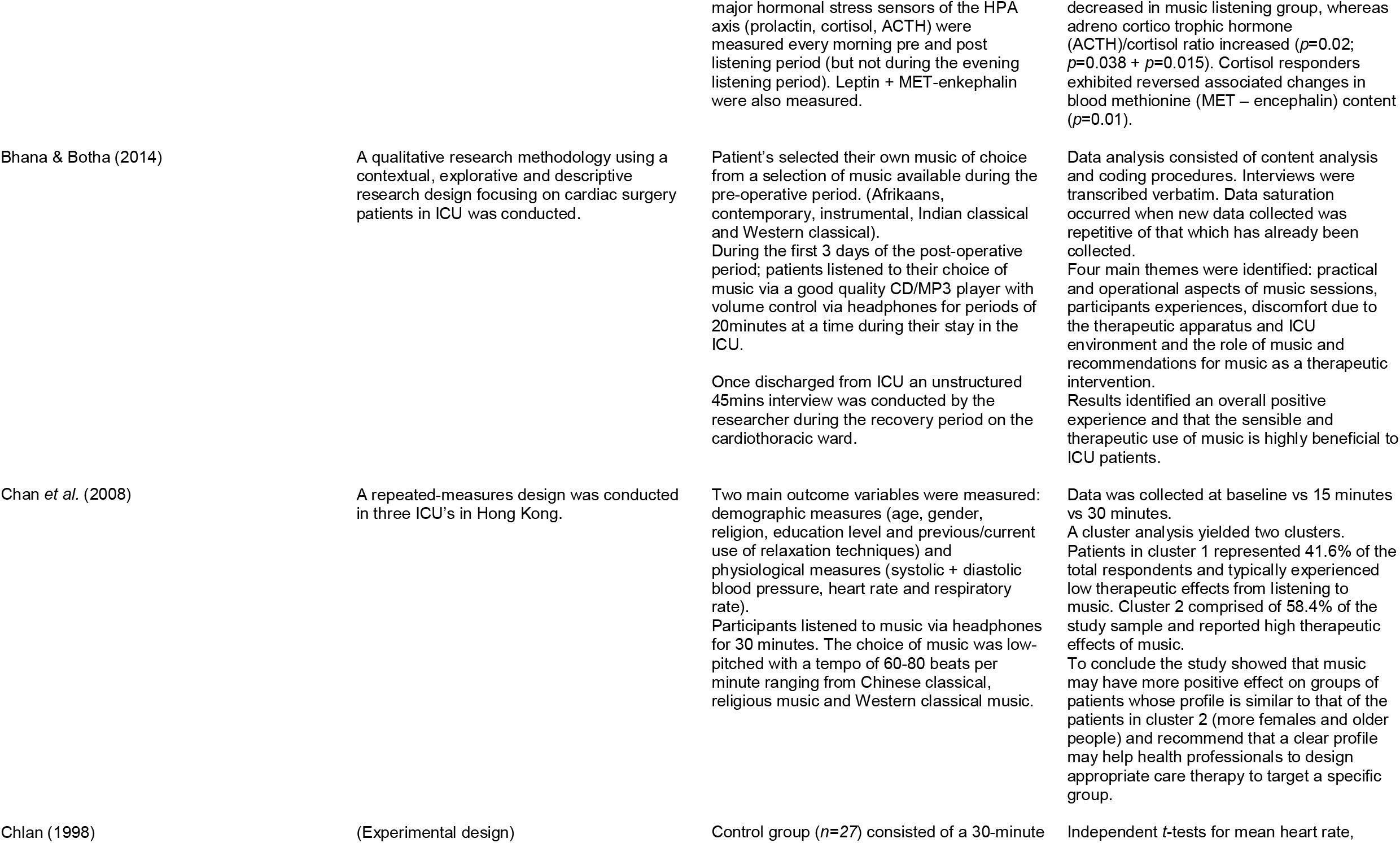
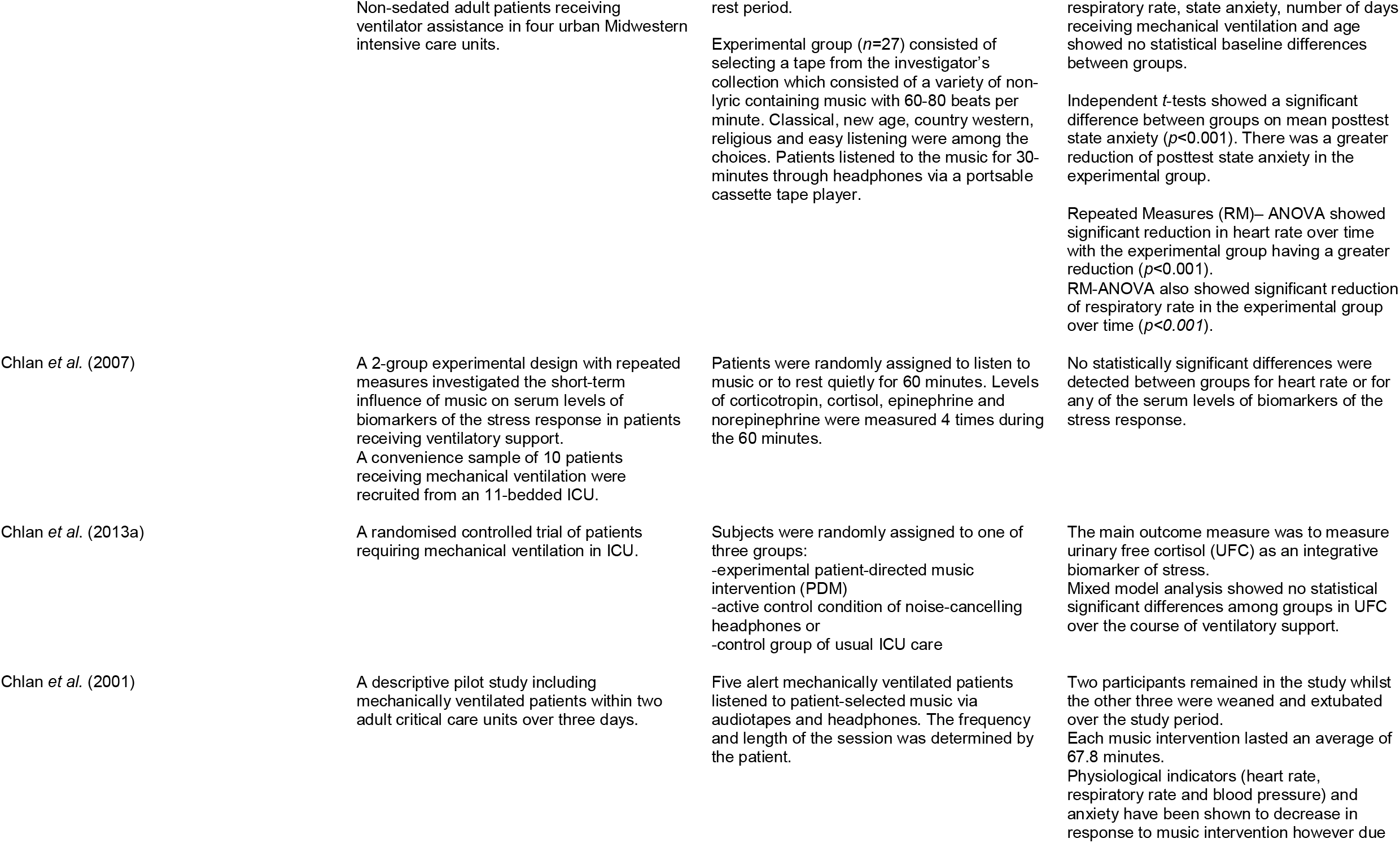
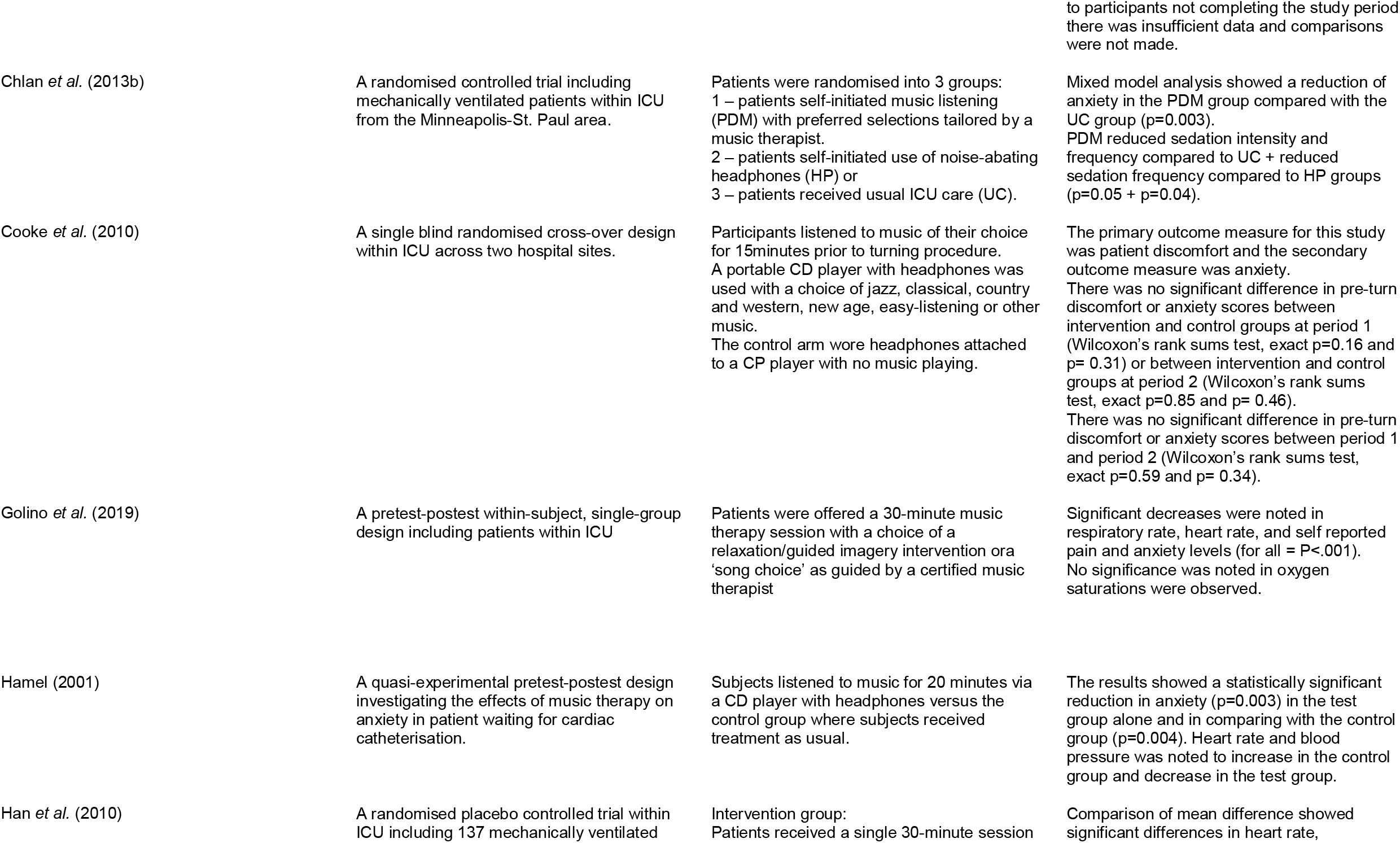
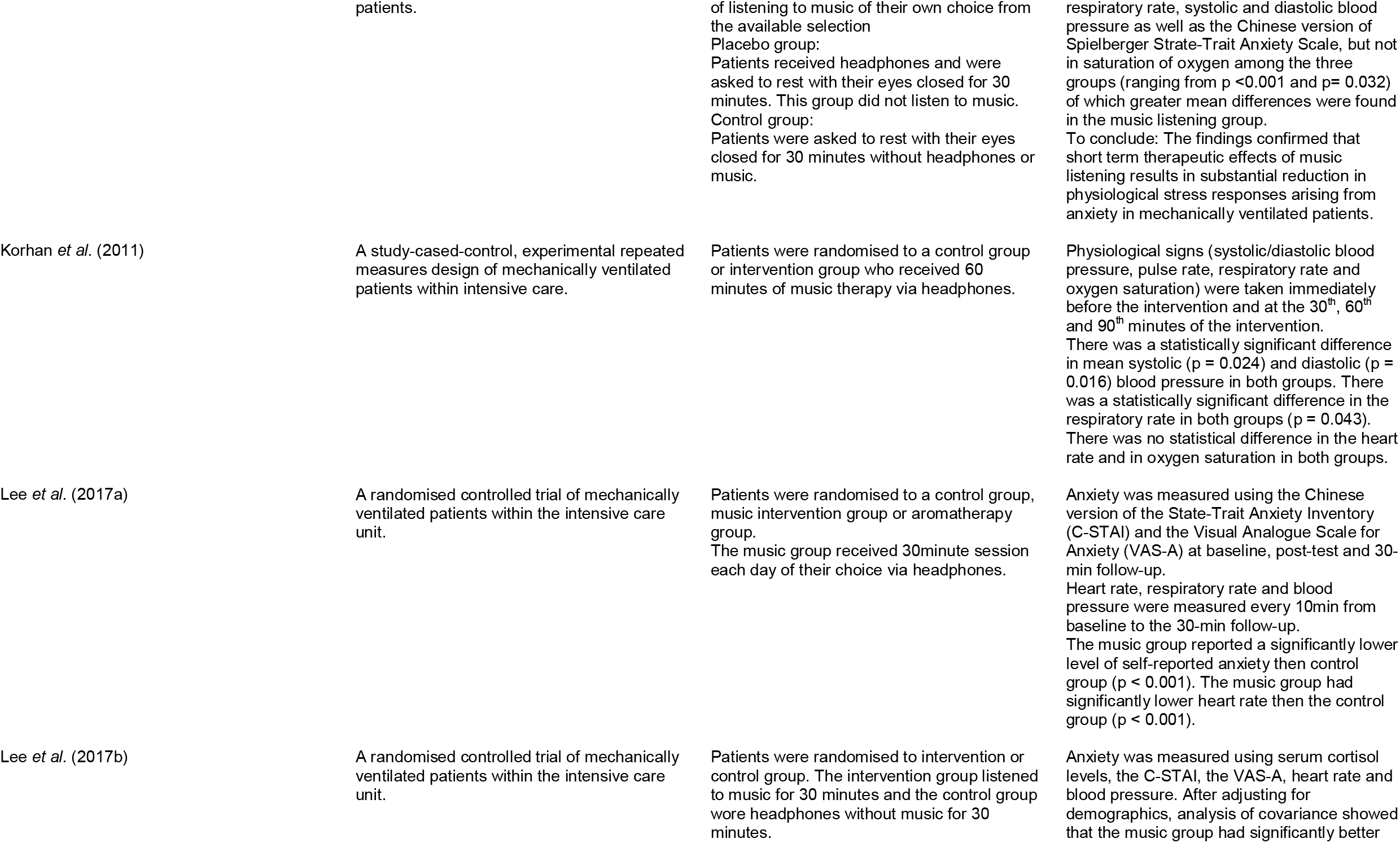
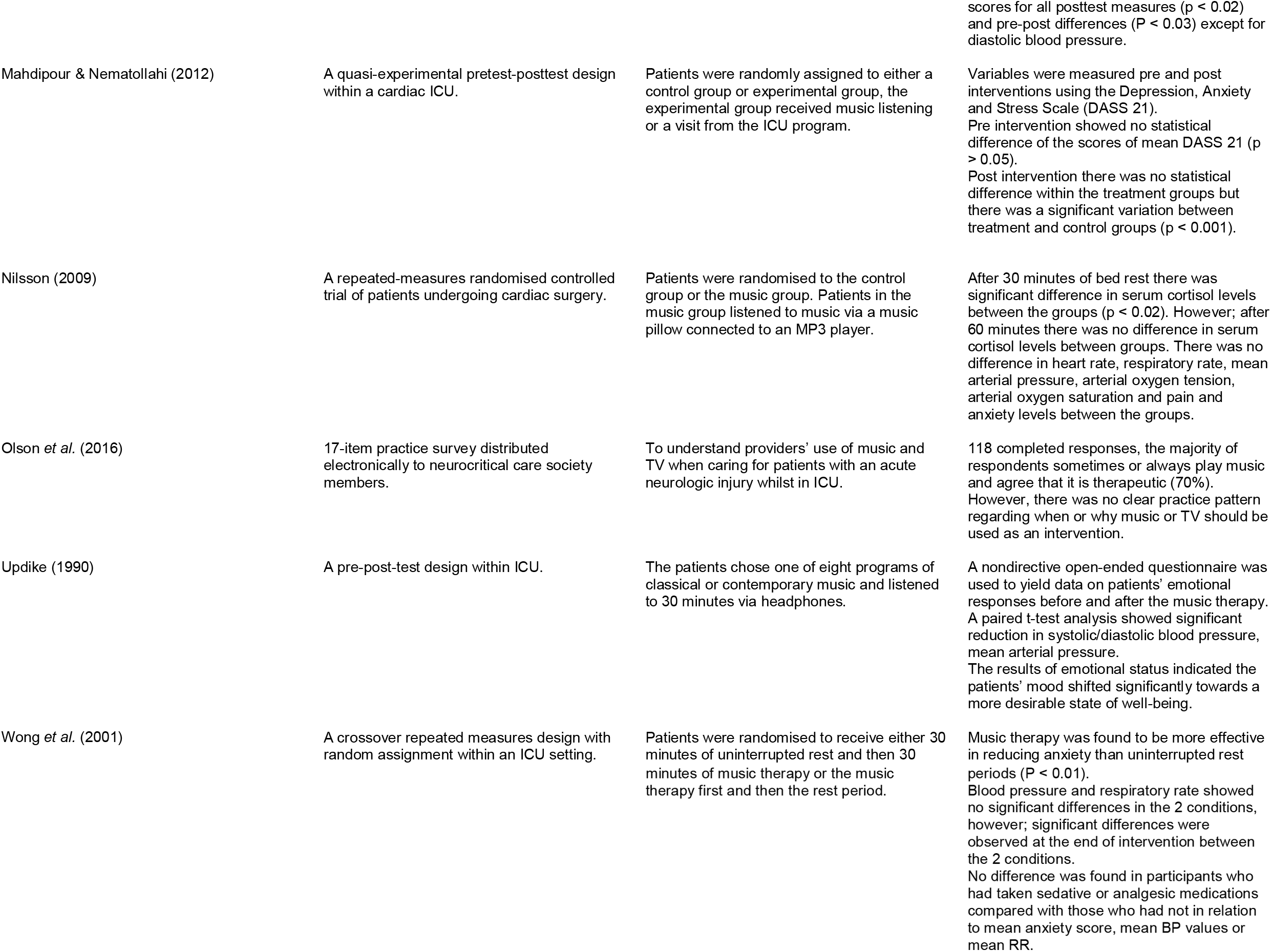
CMO Configurations.

**Table 2.**
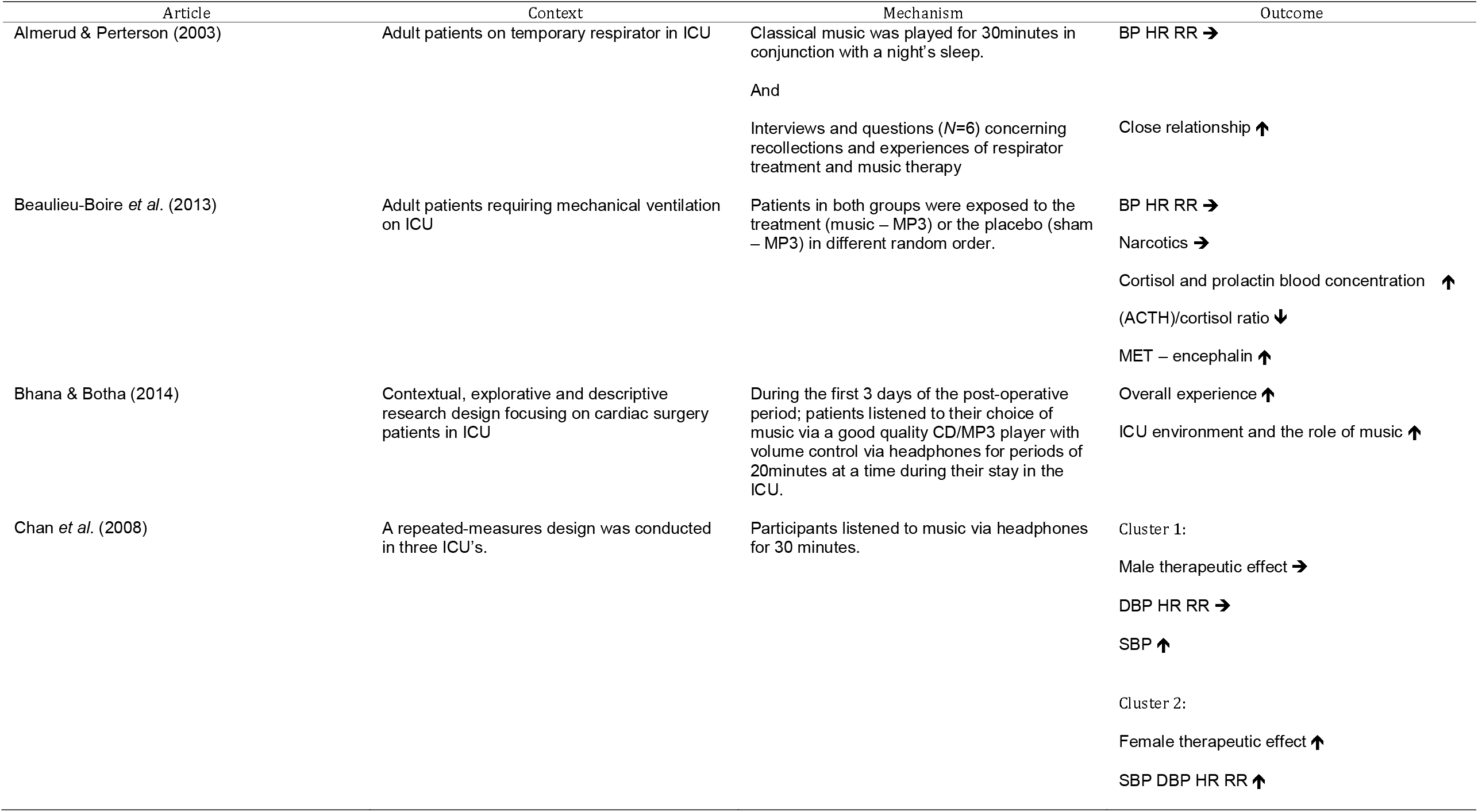
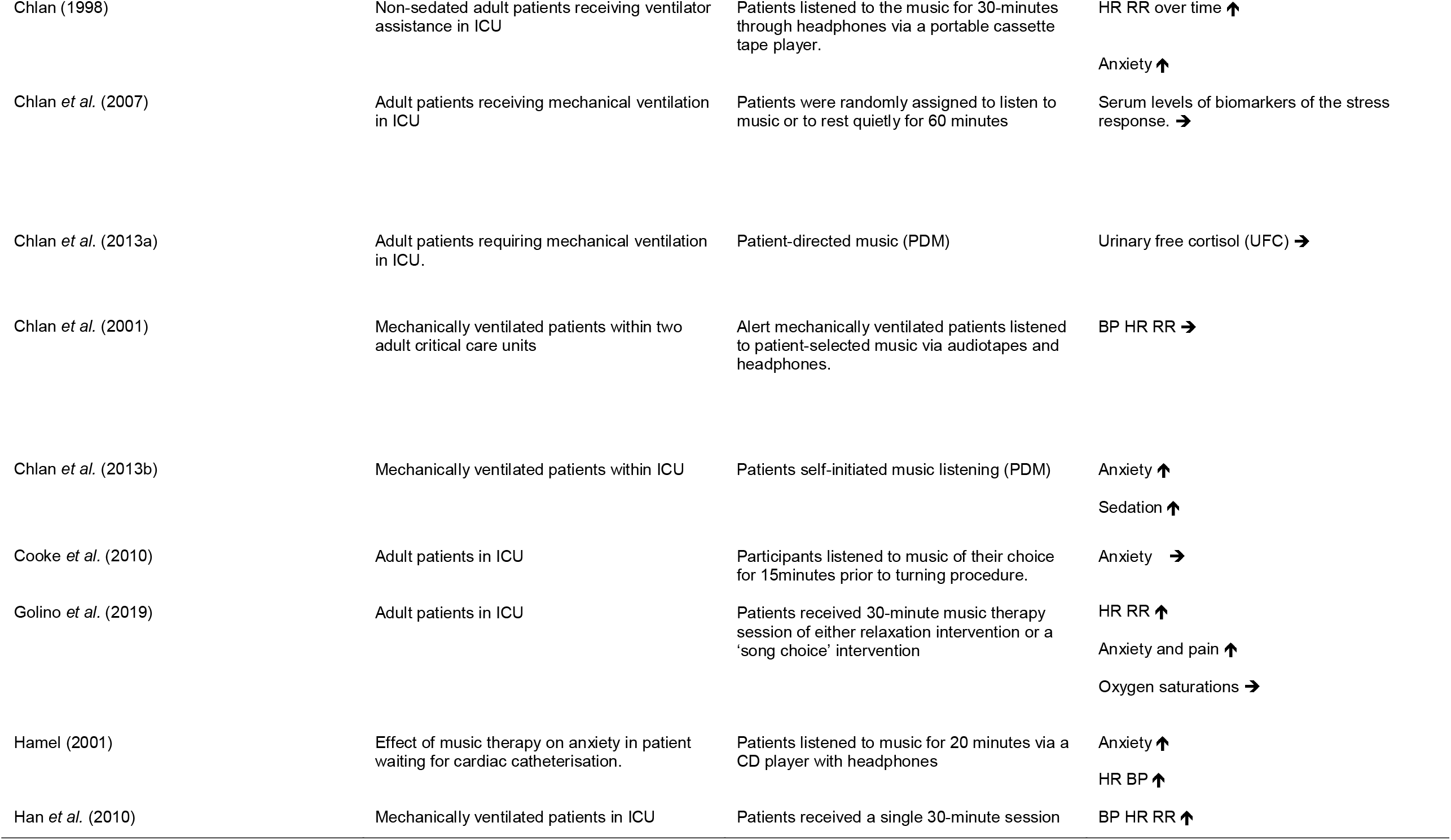
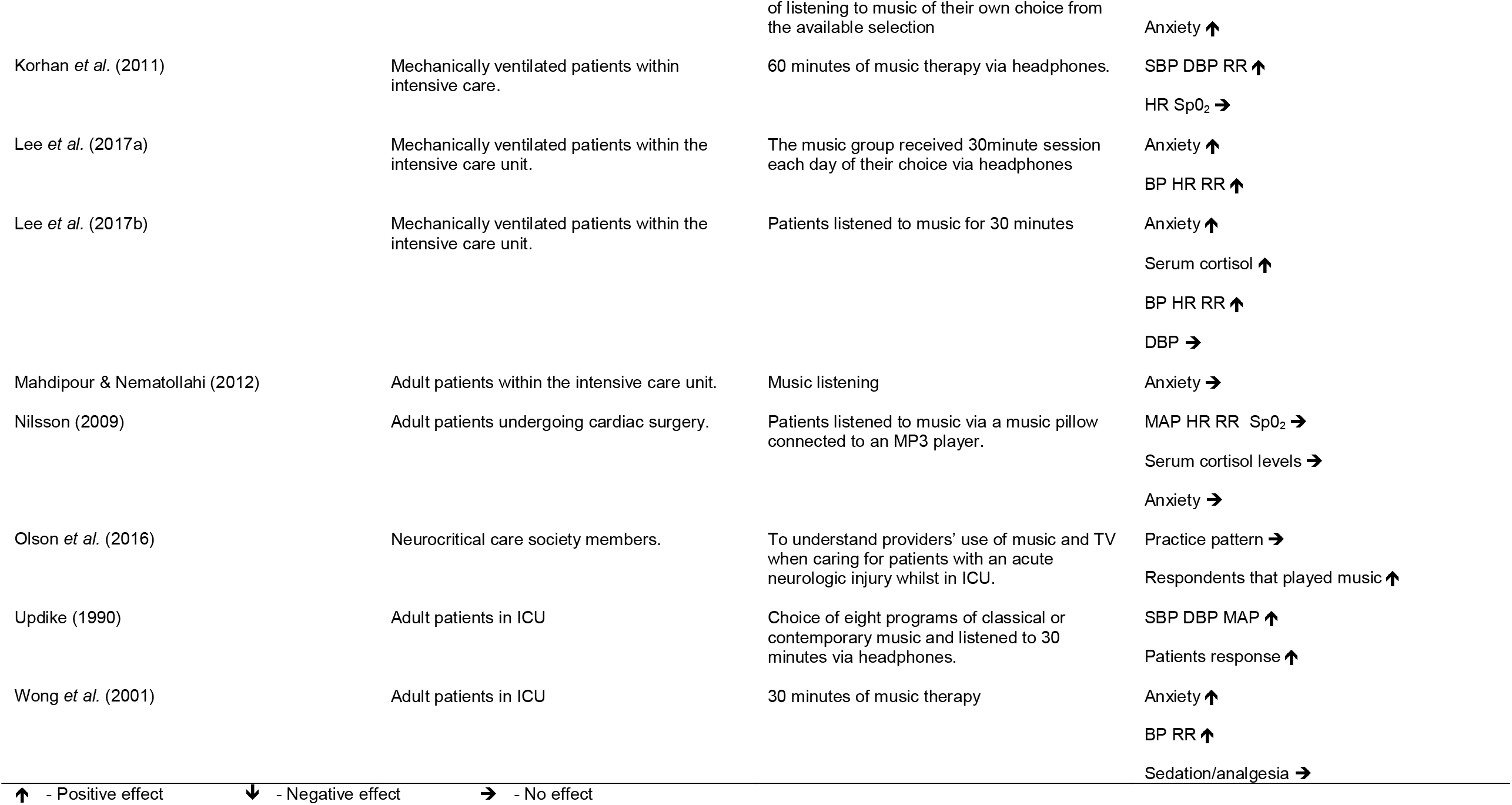
CMO configurations at a glance

### Contexts

Sixteen of the 21 articles included in this review the main context were adult mechanically ventilated patients within ICU (Almedured & Petersson, 2003; Beaulieu-Boire *et al*. 2013; Chan *et al*. 2008; Chlan, 1998; Chlan *et al*. 2007; Chlan *et al*. 2013a; Chlan *et al*. 2013b; Chlan *et al*. 2001; Cooke *et al*. 2010; Han *et al*. 2010; Korhan *et al*. 2011; Lee *et al*. 2017a; Lee, *et al*. 2017b; Maddpour & Nematollahi, 2012; Updike, 1990; Wong *et al*. 2001).

Five of the articles did not focus on mechanical ventilation, Bhana and Botha 2014) and Nilsson (2009) focused on patients undergoing cardiac surgery in ICU and Hamel (2001) focused on patients undergoing cardiac catheterisation. Golino *et al*. (2019) did focus on adult patients in ICU but had excluded mechanically ventilated patients. Oslon *et al* (2016) focused on providers use within neurocritical care.

### Mechanisms

The length of the music sessions ranged from 30 to 60 minutes, and all delivered music via headphones using MP3, cassette or CD player barring Golino *et al*. (2019) who focused their study on the value of active music. Patient directed music was a commonality noted; however, it was from a list chosen by the researchers or a trained music therapist often resulting in a slow tempo and typically relaxing music.

### Outcomes

Anxiety as a phenomenon can be measured in diverse ways as demonstrated throughout the literature and within the context of critically ill mechanically ventilated patients; demi regularities emerged from the data, which identified physiological measurements and state-trait anxiety as the most common outcome measurement (Tables 1 & 2). Thirteen of the 21 studies measured physiological measurements as a marker of anxiety, nine studies noted music had a positive effect on the outcome whereas six of the studies found no effect. Three studies had mixed results; Lee *et al*. (2017b) found a positive effect on BP HR RR but no effect on DBP, Korhan *et al*. (2011) noted a positive effect in SBP, DBP and RR but no effect in HR and Sp0_2_ and Chan *et al*. (2008) found a positive effect on SBP, DBP, HR and RR in cluster 2 but no effect on DBP, HR, RR in cluster 1 and a positive effect in SBP; furthermore, cluster 1 interestingly was male dominant compared with cluster 2. This was the only study to note a difference between male and female response suggesting the latter had a more positive effect from music compared with the former.

Fifteen of the studies measured anxiety using validated tools and found that music had a positive effect on the reduction of anxiety; however, three of the 15 studies found no effect in the reduction of anxiety. Biomarkers of stress were measured in five studies and two of the studies found no effect in biomarkers and music therapy. Lee *et al*. (2017b) found a positive effect in serum cortisol level and Beaulieu-Boire *et al*. (2013) noted a positive effect in cortisol, prolactin and MET-encephalin levels but no effect in ACTH/cortisol ratio. It is important to note that there is heterogeneity amongst the studies and not one study focused on the same biomarkers of stress.

Patient and health care professional’s experiences of music therapy were measured in four studies and all found a positive effect on patients experiences and the role of music in an ICU setting; furthermore, Olson *et al*. (2016) found a positive effect on practitioners playing music. Music was also noted to have a positive effect on sedation in the study by Chlan *et al*. (2013b). Beaulieu-Boire *et al*. (2013) and Wong *et al*. (2001) found no effect on music therapy and analgesia or sedative use.

The overarching results demonstrated that music did not have a negative effect on the outcomes measured suggesting that this is not a harmful mechanism given the context; however, as discussed earlier the quality of evidence is heterogeneous along with the measured outcomes and the mechanisms that lead to the outcomes which does not provide with unambiguous results or a regular pattern.

## DISCUSSION

### What works for whom and under what circumstances?

In keeping with the RRR method, it was difficult to ascertain what works for whom under what circumstances due to the limitations and heterogeneity of the data. The CMO’s were teased from the data, which has concluded that the mechanisms and the outcomes are very poorly agreed in the field and more research is needed. It is clear that music can have a place within critical care and there are some suggestions that the most common mechanism is patient directed music for 30 minutes via headphones, which had a positive effect on the outcome. Overall anxiety was reduced, and music is noted to have a positive effect and additionally can do no harm to the patients whether they are receiving mechanical ventilation or not.

### Implications and recommendations

- Music is a cost effective and can be a nurse lead intervention, therefore; a development of a protocol for nurses to follow could be implemented into practice
- A discussion with the patients’ relatives and the patient if they are conscious could be encouraged by the ICU team to establish whether or not music is important to the patient thus arrangements of bringing in the patient’s own music with headphones can take place
- It is prudent to note that ICU delirium is prevalent within this population of patients. Music may inhibit the recovery process of those suffering with delirium, therefore; regular CAM-ICU testing should be considered and included into the protocol (Barr *et al*. 2013)

## Limitations

- One potential non-English article was not included in the review due to a lack of resources available for translation
- One reviewer completing the review, which can lead to errors in the search process, study selection, data analysis and data interpretation.
- Confirmation bias may have occurred but a reflective approach using peers has been used to check for this
- Bias may have also occurred due to the reviewer being an experienced ICU nurse.
- Limited time and resources to complete comprehensive realist synthesis
- Quality of the review may be limited due to the reviewers lack of experience with the RRR methodology

## Conclusions

Adopting the realist approach enabled the horizons to be broadened in a topic which cannot be quantified alone as it requires the response, thoughts, feelings and stories from patients to enable a deeper understanding of music as a phenomenon within the critical care setting.

What works for whom and under what circumstances? The answer to this question was contradicted throughout the research as not one paper approached the CMO’s the same. Theories could be developed and implemented into a protocol for practice; however, it would be based on the researcher’s own experience alongside poor-quality heterogeneous evidence. More research is required with agreed interventions and outcomes with a mixture of qualitative and quantitative evidence, as the complex nature of music as an intervention and humans as a species, our understanding of music as a therapy remains theoretical and very much a personal commodity.

### Future research

Due to the heterogeneity of the evidence, there is scope to conduct further consistent and robust research to enable the development of a protocol that can be applied in practice. Specifically, there is a need for greater definition and agreement around standardized music interventions. Likewise, there is a need for agreed outcomes to measure the effectiveness of these interventions. Finally, as with so many studies in this area, it is essential that attention is paid to the aspects of design including: sample size; and suitable control and comparison groups.

## Data Availability

The data are available from the author

---

**Figure 1.**
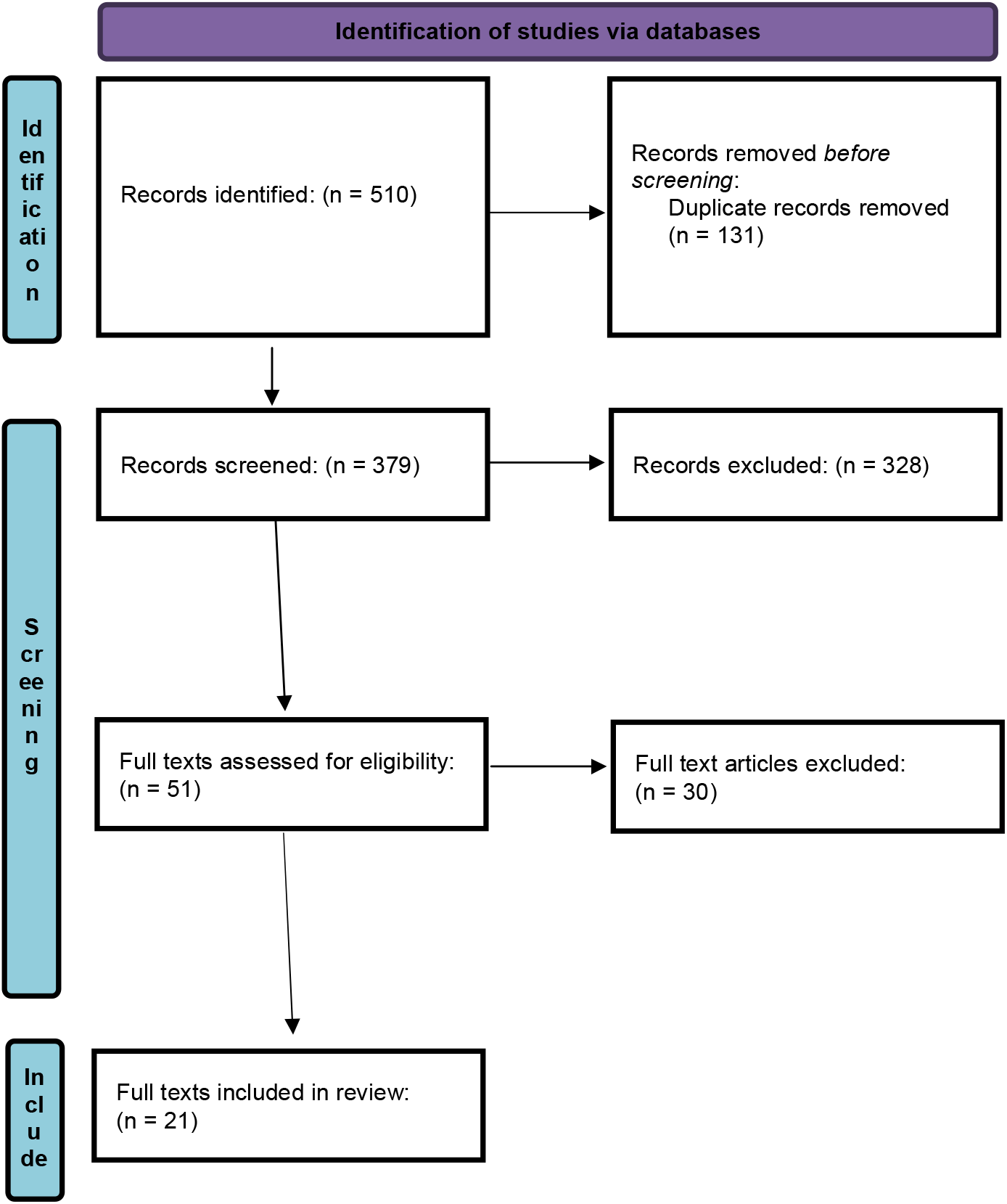
PRIMSA flow diagram.

## Notes

### Competing Interest Statement

The authors have declared no competing interest.

### Funding Statement

No funding was received

### Author Declarations

IRB approval not required

